# Derivation of an Outcome-Driven Threshold for Aortic Pulse Wave Velocity: An Individual-Participant Meta-Analysis

**DOI:** 10.1101/2023.06.06.23291062

**Authors:** De-Wei An, Tine W. Hansen, Lucas S. Aparicio, Babangida Chori, Qi-Fang Huang, Fang-Fei Wei, Yi-Bang Cheng, Yu-Ling Yu, Chang-Sheng Sheng, Natasza Gilis-Malinowska, José Boggia, Wiktoria Wojciechowska, Teemu J. Niiranen, Valérie Tikhonoff, Edoardo Casiglia, Krzysztof Narkiewicz, Katarzyna Stolarz-Skrzypek, Kalina Kawecka-Jaszcz, Antti M. Jula, Wen-Yi Yang, Angela J. Woodiwiss, Jan Filipovský, Ji-Guang Wang, Marek W. Rajzer, Peter Verhamme, Tim S. Nawrot, Jan A. Staessen, Yan Li, The International Database of Central Arterial Properties for Risk Stratification Investigators

## Abstract

**BACKGROUND:** Aortic pulse wave velocity (PWV) predicts cardiovascular events (CVE) and total mortality (TM), but previous studies proposing actionable PWV thresholds have limited generalizability. This individual-participant meta-analysis is aimed at defining, testing calibration, and validating an outcome-driven threshold for PWV, using two populations studies, respectively, for derivation (IDCARS) and replication (MONICA).

**METHODS:** A risk-carrying PWV threshold for CVE and TM was defined by multivariable Cox regression, using stepwise increasing PWV thresholds and by determining the threshold yielding a 5-year risk equivalent with systolic blood pressure of 140 mmHg. The predictive performance of the PWV threshold was assessed by computing the integrated discrimination improvement (IDI) and the net reclassification improvement (NRI).

**RESULTS:** In well-calibrated models in IDCARS, the risk-carrying PWV thresholds converged at 9 m/s (10 m/s considering the anatomical pulse wave travel distance). With full adjustments applied, the threshold predicted CVE (HR [CI]: 1.68 [1.15-2.45]) and TM (1.61 [1.01-2.55]) in IDCARS and in MONICA (1.40 [1.09-1.79] and 1.55 [1.23-1.95]). In IDCARS and MONICA, the predictive accuracy of the threshold for both endpoints was ∼0.75. IDI was significant for TM in IDCARS and for both TM and CVE in MONICA, whereas NRI was not for any outcome.

**CONCLUSIONS:** PWV integrates multiple risk factors into a single variable and might replace a large panel of traditional risk factors. Exceeding the outcome-driven PWV threshold should motivate clinicians to stringent management of risk factors, in particular hypertension, which over a person’s lifetime causes stiffening of the elastic arteries as waypoint to CVE and death.

## INTRODUCTION

Over the human lifespan, aging and age-related risk factors, such as hypertension and type-2 diabetes, lead to stiffening of the central elastic arteries. Consequently, the systolic load on the arterial walls is cushioned less, a phenomenon further amplified by the early return of reflected waves in late systole, while the tensile force maintaining a continuous blood flow during diastole diminishes.^1^ Aortic pulse wave velocity (PWV) is the gold standard for the non-invasive assessment of central arterial stiffness^2^ and predicts adverse cardiovascular (CV) outcomes in a continuous manner.^1^ However, in support of clinical decision making European^3^ and Chinese^4^ guidelines proposed a fixed risk threshold of >12 m/s, which European experts subsequently lowered to >10 m/s, considering the difference between the measured and anatomical pulse wave travel distance.^2,5-7^

The literature supporting the guideline-endorsed PWV thresholds^2-5^ consists of patient^8-15^ and community-based^16-19^ studies with total or CV mortality or a composite CV endpoint as outcome. In patients with end-stage kidney disease,^9-12,15^ metabolic syndrome,^14^ hypertension,^8^ or in patients undergoing transluminal aortic valve replacement,^13^ PWV risk thresholds ranged from 10.5 m/s^11^ to 11.8 m/s.^12^ In Japanese-Americans,^16^ Japanese men,^20^ or middle-aged or elderly community-dwelling individuals,^17,19^ the PWV risk thresholds ranged from 9.0 m/s^18^ to 13.7 m/s.^19^ In a cross-sectional meta-analysis of 16,867 individuals,^7^ the distribution of PWV was described as function of age in various patient strata, including a subset of 1455 patients with optimal or normal blood pressure (BP) considered to mirror population-based levels. Notwithstanding the merits of the previous publications,^11-14,16-19^ PWV thresholds derived in patients with advanced disease^11-13^ or disturbed metabolic profile,^14^ in single-center population cohorts,^16-19^ in the elderly,^17^ or based on the PWV distribution rather than adverse health outcomes^7^ cannot be straightforwardly extrapolated to clinical practice. In view of these potential limitations, the current individual-participant level meta-analysis, covering a wide age range, was prospectively and specifically designed to define, test the calibration, and validate an outcome-driven threshold for PWV, using the International <u>D</u>atabase of <u>C</u>entral <u>A</u>rterial Properties for <u>R</u>isk <u>S</u>tratification (IDCARS)^21^ as derivation dataset and the Copenhagen subset of the <u>Moni</u>toring of Trends and Determinants in <u>Ca</u>rdiovascular Disease Study (MONICA) for replication.^22^

## METHODS

### Data Availability

All available data are shown within the article and the online-only Data Supplement. Anonymized data are available from the corresponding author upon request, on condition that an analysis plan is accompanying the request and that the principal investigators of all cohorts approve data sharing.

### Study Cohorts

The population studies included in the current meta-analysis met the principles outlined in the Helsinki declaration for investigation of human participants.^23^ The IDCARS study protocols and the secondary analyses of anonymized data were approved by the competent local Institutional or National Review Boards. Anonymized data from the Copenhagen subset of the MONICA study were used for analysis. Participants gave written informed consent at recruitment and renewed consent at each follow-up visit. The online only Data Supplement provides full details on the selection of the study population and the methods applied for collecting the clinical, biochemical and hemodynamic measurements and the statistical analysis.

#### IDCARS — Derivation Cohort

IDCARS cohorts qualified for inclusion in the present analysis, if peripheral and central BP and CV risk factors had been measured at baseline, and if follow-up included both fatal and nonfatal outcomes. Eight cohorts met these eligibility criteria (**Table S1**). Initial enrollment took place from 1985 until 2015. For the present analysis, baseline refers to the first measurement of central and peripheral BP along with CV risk factors (October 2000 until February 2016). Across studies, the last follow-up took place from October 2012 to December 2018 (**Table S1**). In the 8 qualifying IDCARS cohorts, 6546 individuals took part in a re-examination including also the vascular examination. Of those, 2706 (41.3%) only underwent a tonometric pulse wave analysis or had a substandard assessment of PWV. Of the remaining 3840 participants, 462 (12.0%) were discarded, because they were younger than 30 years, leaving 3378 IDCARS participants for statistical analysis.

#### MONICA — Replication Cohort

In 1982-1984, a random sample of the residents of Glostrup County, one of the Western suburbs of Copenhagen, Denmark was drawn with the goal to recruit an equal number of women and men aged 30, 40, 50, and 60 years. In 1993-1994, the 3785 former participants were invited for a follow-up examination at the Research Center for Prevention and Health in Glostrup, of whom 2493 (65.9%) without history of CV disease between recruitment and follow-up were examined.^22^ For the current analysis, 35 (1.40%) were excluded because of inaccurate or missing PWV measurements, leaving 2458 participants for analysis.

### BP and PWV Measurement

In IDCARS, brachial BP was the average of the first 2 consecutive readings. Mean arterial pressure (MAP) was peripheral diastolic BP plus one-third of pulse pressure. In all IDCARS cohorts included in the current analysis, PWV was measured by sequential electrocardiographically gated recordings of the arterial pressure waveform at the carotid and femoral arteries. The observers measured the distance from the suprasternal notch to the carotid sampling site (distance A), and from the suprasternal notch to the femoral sampling site (distance B). Pulse wave travel distance was calculated as distance B minus distance A.^21^ Pulse transit time was the average of 10 consecutive beats.^21^ PWV is the ratio of the travel distance in meters to transit time in seconds. PWV measurements were discarded if the standard error of the mean of 10 beats was more than 10%.^21^

In MONICA, a trained nurse obtained 2 consecutive BP readings with a random zero mercury sphygmomanometer, which were averaged for analysis. Immediately thereafter, the trained nurse used 2 piezoelectric pressure transducers (Hellige GmbH, Freiburg im Breisgau, Germany) to record the arterial wave simultaneously at the left common carotid and femoral arteries. ^24^ PWV was the travel distance between the 2 transducers, measured on the body surface, divided by the transit time, determined manually by the foot-to-foot velocity method.^24^ For analysis, PWV measurements from 2 to 15 heart cycles were averaged. The directly measured path length (MONICA) was converted to the subtracted path length (IDCARS) in analyses involving both cohorts.^7,25^ For comparison of IDCARS and MONICA data in normal participants with a published meta-analysis,^7^ path length was converted to the path length considered to reflect the true anatomical distance, using published formula.^7^ The biochemical methods are available in the online-only Data Supplement (**p S5**).

### Ascertainment of Endpoint

The co-primary endpoints in the current study were a composite CV endpoint, including CV death and nonfatal CV events, and total mortality. The secondary endpoints included CV mortality and fatal combined with nonfatal coronary events are defined in the Data Supplement (pp **S5-S6**). In all outcome analyses, only the first event within each category was considered. Participants free of events were censored at last follow-up.

### Statistical Analysis

Statistical methods are described in detail in the online-only Data Supplement (pp **S6-S9**). In exploratory analyses, incidence rates of endpoints were tabulated by tertiles of the PWV distribution, while applying the direct method for standardizing rates in IDCARS for cohort, sex and age (<40, 40-59, ≥60 years) and for sex and age group (40, 50, 60, and 70 years) in MONICA. The cumulative incidence of the primary and secondary endpoints was plotted, while accounting for cohort (in IDCARS only) and sex and age (IDCARS) or age group (MONICA).

Multivariable-adjusted Cox models accounted for sex, age (IDCARS) or age group (MONICA), MAP, heart rate, body mass index, the total-to-HDL serum cholesterol ratio, smoking and drinking, use of antihypertensive drugs, history of diabetes mellitus,^26^ and previous CV disease (IDCARS only). Multivariable analyses involving IDCARS additionally accounted for cohort. The proportional hazards assumption was checked by the Kolmogorov-type supremum test. To compare relative risk across strata, deviation from mean coding^27^ was applied.

To determine an operational threshold for PWV, a two-pronged strategy^28,29^ was applied using Cox regression in IDCARS. First, multivariable-adjusted HRs were computed for 0.1 m/s increments in PWV from the 10th to the 90th percentile of the PWV distribution. These HRs expressed the risk in participants, whose PWV exceeded the cut-off point *vs* the average risk in the whole population. The HRs with CIs were plotted as function of increasing PWV thresholds to assess at which PWV level the lower 95% confidence limit of the HRs crossed unity, signifying increased risk.^28^ Next, PWV thresholds were obtained by determining the PWV levels yielding a 5-year risk equivalent to the risk associated with an office systolic BP of 120-, 130-, 140- and 160 mm Hg.^29^ Model calibration was evaluated by comparing the predicted risk against overoptimism-corrected Kaplan-Meier estimates in PWV quintiles. The performance of PWV in risk stratification was assessed from 2-by-2 tables providing specificity, sensitivity and related statistics, the area under the curve, and by the integrated discrimination improvement (IDI) and the net reclassification improvement (NRI).^30^ Finally, subgroup analyses were conducted in participants stratified by sex, age (<60 *vs* ≥60 years), and the approximate median systolic BP in IDCARS and MONICA (<130 *vs* ≥130 mm Hg).

## RESULTS

### Baseline Characteristics

In IDCARS, the number of interpolated values amounted to 108 (3.2%) for total serum cholesterol, 198 (5.9%) for HDL cholesterol, 89 (2.6%) for blood glucose, 161 (4.8%) for smoking and 723 (21.4%) for use of alcohol. In MONICA, the corresponding number of interpolated values were 1 (0.04%), 1 (0.04%), 5 (0.20%), 0 (0%), and 49 (1.99%), respectively.

The differences in the baseline characteristics between the IDCARS (2000-2016) and the MONICA (1993-1994) participants at the time of the vascular examinations (**Table 1**) reflect how lifestyle and treatment rates of hypertension changed over time, the ethnic make-up and the age structure of the discovery and replication cohorts, and the high tax rates levels levied on alcoholic beverages in Denmark. In both IDCARS and MONICA, women compared with men had smaller body height, lower body weight and body mass index, lower systolic and diastolic BP and MAP, and higher HDL cholesterol (**Table S2**). Across the IDCARS cohorts, mean PWV (SD) ranged from 7.2 (1.5) m/s in the Polish Gdańsk cohort to 8.9 (2.4) m/s in the participants recruited in Buenos Aires, Argentina (**Figure S1**). In the Copenhagen MONICA cohort, PWV averaged 8.0 (2.5) m/s (**Figure S1**).

**Table 1.**
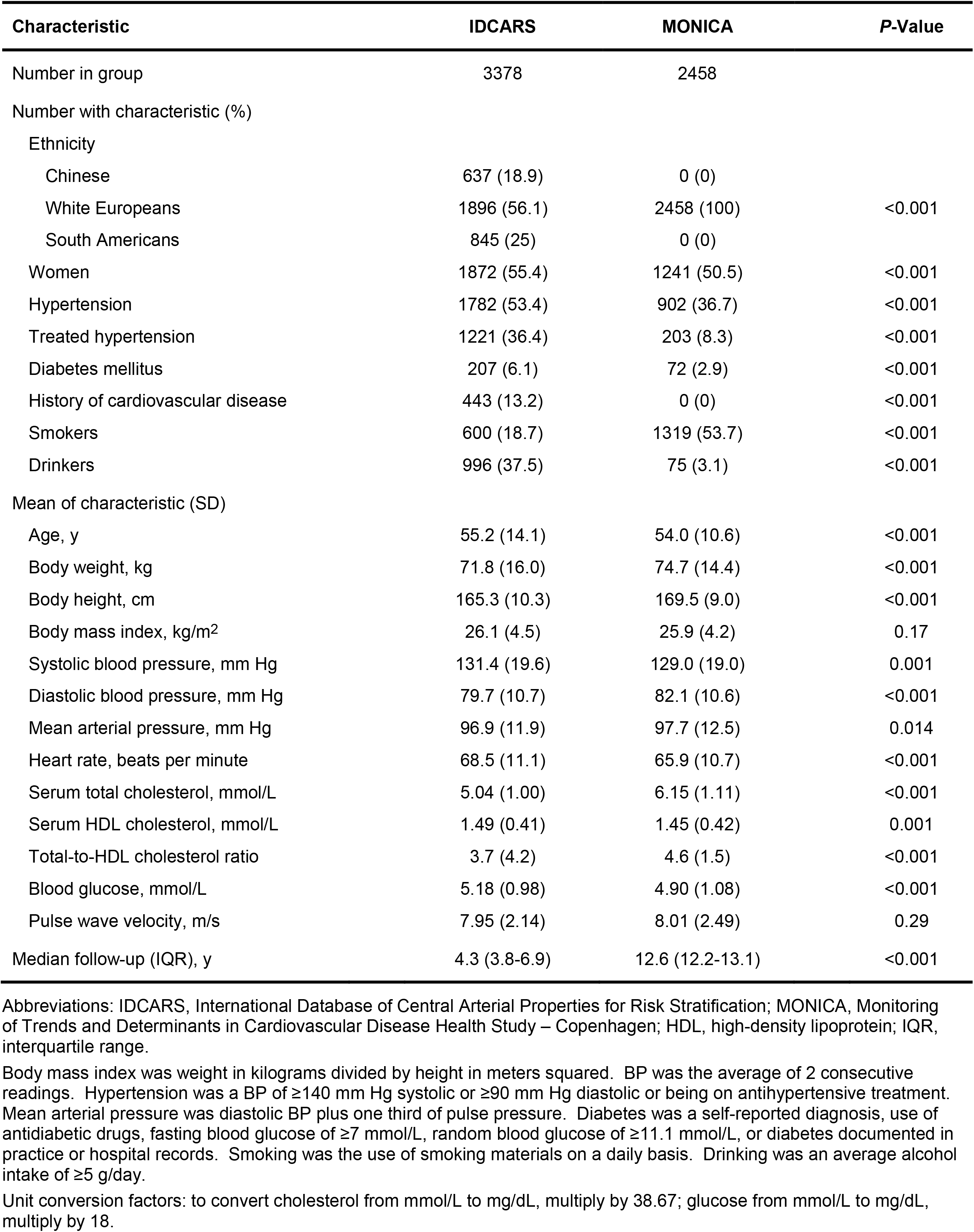
Baseline Characteristics by Cohort.

To assess concordance between the data resources used, normal individuals were sampled from IDCARS and MONICA by excluding patients with a history of CV disease, diabetes or treated hypertension and by discarding patients with dyslipidemia and smokers (**Figure S2**). With standardization of the travel path applied, PWV by age group was largely similar in IDCARS and MONICA (**Table S3**). Furthermore, in single regression, the main correlates of PWV were age, systolic BP, pulse pressure and MAP (**Table S4**) with Pearson correlation coefficients amounting to were 0.57, 0.45, 0.43 and 0.33 in IDCARS and to 0.55, 0.53, 0.48 and 0.47 in MONICA (*P*<0.001 for all).

### Incidence of Endpoints

Median follow-up of IDCARS participants amounted to 4.3 years (IQR: 3.8-6.9 years; 5th-95th percentile interval: 2.1-11.6 years. Over follow-up (**Table S5**), 105 participants (3.11%) died: 25 (0.74%) because of CV disease, 74 (2.19%) because of a non-CV illness (including kidney failure), and 6 (0.18%) because of non-documented illnesses. The number of IDCARS participants experiencing a major CV event or a coronary endpoint amounted to 155 (4.59%) and 77 (2.28%), respectively.

Median follow-up in MONICA was 12.6 years (IQR: 12.2-13.1 years; 5th to 95th percentile interval: 3.5-13.4 years). Over this time period (**Table S5**), 393 (16.0%) died: 139 (5.66%) because of CV disease and 254 (10.3%) because of a non-CV ailment. The number of MONICA participants experiencing a major CV event, or a coronary endpoint amounted to 354 (14.4%) and 202 (8.22%), respectively. Across tertiles of the PWV distribution, in IDCARS as well as in MONICA, the co-primary (**Table S6**) and secondary (**Table S7**) endpoints steeply increased with higher PWV category (*P*<0.001).

### PWV Thresholds in IDCARS

Multivariable-adjusted HRs were plotted against PWV thresholds stepwise increasing by 0.1-m/s over the 10th-90th percentile range of the PWV distribution (**Figure 1A** and **Figure 1B**). These multivariable-adjusted HRs expressed the 5-year risks of the co-primary endpoints associated with successively increasing PWV thresholds compared to the average risk in the whole IDCARS cohort. The lower limit of the 95% CI of the HRs crossed unity at a PWV level of 8.7 m/s for the composite CV endpoint and at 8.8 m/s for total mortality. In multivariable-adjusted Cox models (**Figure 1C** and **Figure 1D**), the PWV thresholds yielding a risk equivalent with a systolic BP of 140 mm Hg were 8.5 m/s (CI: 7.5-9.6) for the composite CV endpoint and 8.2 m/s (7.1-9.4) for total mortality. In all models PWV met the proportional hazard assumption (test statistic ≤1.26; *P*≥0.175). In IDCARS, with adjustments applied for cohort, sex, age and MAP, a PWV threshold of <9 m/s *vs* ≥9 m/s separated (**Figure S3**) the cumulative incidence of the co-primary and secondary endpoints in a highly significantly way (*P*≤0.013). The fully adjusted HRs associated with a PWV ≥9 m/s vs <9 m/s, were 1.68 (CI: 1.15-2.45) for the composite CV endpoint and 1.61 (1.01-2.55) for total mortality (**Table 2**); for CV mortality and coronary endpoints (**Table S8**), the corresponding HRs were 3.21 (1.06-9.69) and 2.08 (1.21-3.59), respectively. In **Table 2** and **Table S8**, the HRs associated with a 1-SD PWV increment were also presented allowing comparison with the literature. The fully adjusted Cox models including the 9-m/s PWV threshold were well calibrated as evidenced by similarity between the Kaplan-Meier estimates and the multivariable-adjusted mean predicted risk for the CV endpoint (*P*=0.654) and total mortality (*P*=0.691) across quintiles of observed and predicted risk (**Figure 1E** and **Figure 1F**). In subgroup analyses stratified for sex, age or median systolic BP, none of the interactions between the 9-m/s threshold and stratification groups reached significance (**Figure S4**).

**Figure 1.**
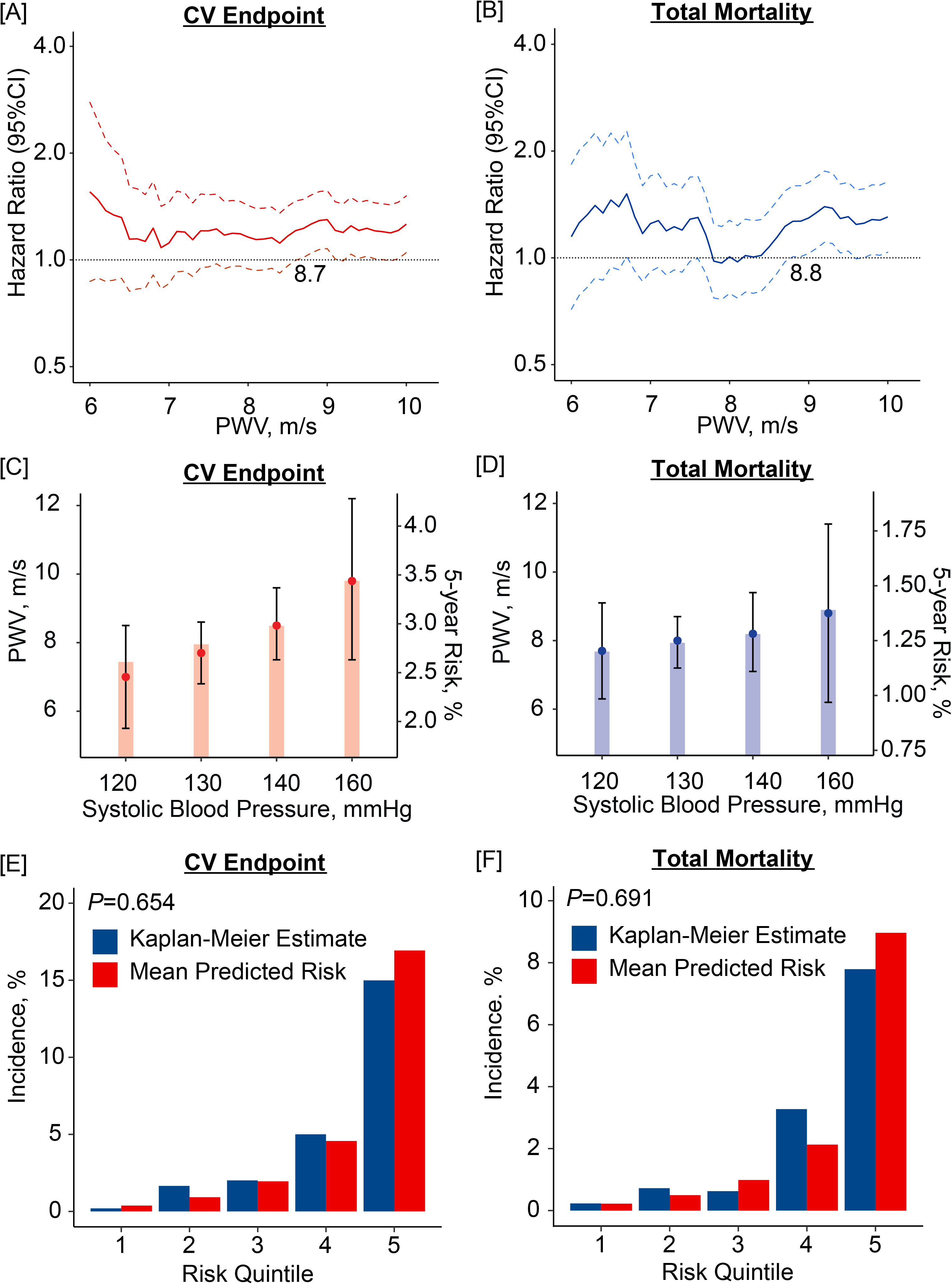
Threshold and Calibration of Pulse Wave Velocity in 3378 IDCARS participants. Hazard ratios (HRs) express the risk at each PWV level relative to the average risk in the whole study population for composite cardiovascular endpoint (**A**) and total mortality (**B**) with PWV at 8.7 and 8.8 m/s signifying increased risk by crossing unity (dotted line). PWV levels yielding equivalent 5-year risks compared with systolic blood pressure categories for composite cardiovascular endpoint (**C**) and total mortality (**D**) with bars indicating 5-year risks and point and line for PWV thresholds. PWV levels at 8.5 and 8.2 m/s indicate equivalent risk as a systolic blood pressure of 140 mmHg. Model calibration for the composite cardiovascular endpoint (**E**) and total mortality (**F**), showing the predicted risk against overoptimism-corrected Kaplan-Meier estimates in PWV quintiles. All analyses were multivariable adjusted for cohort, sex, age, mean arterial pressure (excluding **C** and **D**), heart rate, the total-to-high-density lipoprotein serum cholesterol ratio, smoking and drinking, use of antihypertensive drugs, diabetes, and history of cardiovascular disease.

**Table 2.** Co-Primary Endpoints in Relation to PWV Per Threshold and Analyzed as Continuously Distributed Variable.

| Cohort / Endpoint | Ne/Nr | Model 1 |  | Model 2 |  |
| --- | --- | --- | --- | --- | --- |
|  |  | HR (95% CI) | P-Value | HR (95% CI) | P-Value |
| IDCARS (discovery) |  |  |  |  |  |
| Cardiovascular endpoint |  |  |  |  |  |
| PWV ≥9 vs <9 m/s | 92/807 vs 63/2571 | 1.76 (1.21-2.56) | 0.003 | 1.68 (1.15-2.45) | 0.007 |
| PWV (+1 SD) | 155/3378 | 1.18 (1.03-1.36) | 0.019 | 1.18 (1.02-1.36) | 0.025 |
| Total mortality |  |  |  |  |  |
| PWV ≥9 vs <9 m/s | 57/807 vs 48/2571 | 1.75 (1.10-2.76) | 0.017 | 1.61 (1.01-2.55) | 0.045 |
| PWV (+1 SD) | 105/3378 | 1.36 (1.17-1.58) | <0.001 | 1.32 (1.13-1.55) | <0.001 |
| MONICA (replication) |  |  |  |  |  |
| Cardiovascular endpoint |  |  |  |  |  |
| PWV ≥9 vs <9 m/s | 160/576 vs 194/1882 | 1.56 (1.23-1.98) | <0.001 | 1.40 (1.09-1.79) | 0.008 |
| PWV (+1 SD) | 354/2458 | 1.27 (1.18-1.37) | <0.001 | 1.24 (1.14-1.34) | <0.001 |
| Total mortality |  |  |  |  |  |
| PWV ≥9 vs <9 m/s | 194/576 vs 199/1882 | 1.71 (1.37-2.13) | <0.001 | 1.55 (1.23-1.95) | <0.001 |
| PWV (+1 SD) | 393/2458 | 1.26 (1.17-1.35) | <0.001 | 1.22 (1.13-1.32) | <0.001 |

Ne/Nr indicates number of events/number of participants at risk. PWV in MONICA was standardized to the subtraction method used in IDCARS. Model 1 accounted for cohort (IDCARS only), sex, age (IDCARS) or age group (MONICA), and MAP. Model 2 additionally accounted for the total-to-HDL serum cholesterol ratio, smoking and drinking, use of antihypertensive drugs, and history of CV disease (IDCARS only).

### Replication in MONICA

The MONICA data were interrogated to replicate the clinical relevance of the proposed 9-m/s threshold. In line with the IDCARS findings, with adjustments applied for sex, age group and MAP, a PWV threshold of <9 m/s *vs* ≥9 m/s differentiated (**Figure S5**) the cumulative incidence of the co-primary and secondary endpoints in a significant manner (*P*≤0.001), except for coronary event (*P*=0.145). The fully adjusted HRs associated with a PWV ≥9 m/s *vs* <9 m/s, were 1.40 (1.09-1.79) for the CV endpoint and 1.55 (1.23-1.95) for total mortality (**Table 2**); for CV mortality (**Table S8**) the HR was 1.53 (1.04-2.25), and for coronary events it was not significant (*P*=0.553). In subgroup analyses stratified for sex, age or median systolic BP, none of the interactions between the 9-m/s threshold and the stratification groups reached significance (**Figure S4**).

### Predictive Performance

In IDCARS, specificity, sensitivity and overall accuracy of the categorized PWV for the prediction of the CV endpoint were 0.775, 0.607 and 0.769 and for the prediction of death 0.767, 0.571 and 0.764, respectively. Estimates in MONICA were of similar magnitude (**Table 3**). In IDCARS, IDI for the 9-m/s PWV threshold amounted to 0.59% for the CV endpoint (*P*=0.020) and to 0.28% (*P*=0.198) for total mortality, while in MONICA the corresponding IDI values were 0.47 (*P*=0.028) and 0.85 (*P*=0.002), respectively (**Table 4**). However, none of the NRI estimates in IDCARS or MONICA reached statistical significance (*P*≥0.108).

**Table 3.** Discriminative Performance of Pulse Wave Velocity.

| Outcome | IDCARS | MONICA |
| --- | --- | --- |
| **Composite Cardiovascular Endpoint** |  |  |
| Categorized PWV ($\geq 9$ vs $< 9$ m/s) | | |
| N° events/at risk (%) | 74/807 vs 48/2571 | 125/576 vs 142/1882 |
| Specificity (95% confidence interval) | 0.775 (0.760-0.789) | 0.794 (0.777-0.811) |
| Sensitivity (95% confidence interval) | 0.607 (0.514-0.694) | 0.468 (0.407-0.530) |
| PLR (95% confidence interval) | 2.694 (2.304-3.151) | 2.274 (1.954-2.648) |
| NLR (95% confidence interval) | 0.508 (0.407-0.633) | 0.670 (0.597-0.751) |
| Accuracy | 0.769 (0.754-0.783) | 0.759 (0.741-0.776) |
| AUC (95% confidence interval) | 0.691 (0.647-0.735) | 0.631 (0.600-0.662)* |
| Continuously distributed PWV |  |  |
| N° events/at risk (%) | 122/3378 | 267/2458 |
| AUC (95% confidence interval) | 0.749 (0.705-0.793) | 0.710 (0.679-0.742) |
| **Total Mortality** |  |  |
| Categorized PWV ($\geq 9$ vs $< 9$ m/s) | | |
| N° deaths/at risk (%) | 36/807 vs 27/2571 | 136/576 vs 133/1882 |
| Specificity (95% confidence interval) | 0.767 (0.753-0.782) | 0.799 (0.782-0.816) |
| Sensitivity (95% confidence interval) | 0.571 (0.440-0.695) | 0.506 (0.444-0.567) |
| PLR (95% confidence interval) | 2.457 (1.967-3.070) | 2.515 (2.176-2.907) |
| NLR (95% confidence interval) | 0.558 (0.420-0.743) | 0.619 (0.547-0.700) |
| Accuracy | 0.764 (0.749-0.778) | 0.767 (0.750-0.783) |
| AUC (95% confidence interval) | 0.669 (0.607-0.731) | 0.652 (0.621-0.683) |
| Continuously distributed PWV |  |  |
| N° deaths/at risk (%) | 63/3378 | 269/2458 |
| AUC (95% confidence interval) | 0.714 (0.648-0.779) | 0.724 (0.693-0.756) |

Calculations were performed for the 5-year risk in IDCARS and the 10-year risk in MONICA. PLR is the positive likelihood ratio (true positive rate/false positive rate). NLR is the negative likelihood ratio (false negative rate/true negative rate). Accuracy is the overall probability that an individual is correctly classified. All estimates in this table were unadjusted for other risk factors. Significance of the AUC difference between IDCARS and MONICA: \* $P \leq 0.05$

**Table 4.** Integrated Discrimination Improvement and Net Reclassification Improvement by Adding Pulse Wave Velocity Per Threshold and as Continuously Distributed Variable to the Base Model.

| Cohort<br>Endpoint<br>PWV Marker | Integrated Discrimination Improvement |  |  | Net Reclassification Improvement |  |  |
| --- | --- | --- | --- | --- | --- | --- |
|  | IDI (%) | 95% CI (%) | P-value | NRI (%) | 95% CI (%) | P-Value |
| **IDCARS (discovery)** |  |  |  |  |  |  |
| Cardiovascular endpoint |  |  |  |  |  |  |
| PWV $\geq 9$ vs $< 9$ m/s | 0.59 | (0.01, 1.83) | 0.020 | 8.83 | (-2.39, 21.0) | 0.139 |
| Continuous | 0.52 | (0.01, 1.74) | 0.020 | 2.45 | (-17.5, 13.4) | 0.772 |
| Total mortality |  |  |  |  |  |  |
| PWV $\geq 9$ vs $< 9$ m/s | 0.28 | (-0.17, 1.19) | 0.198 | 13.0 | (-10.5, 25.3) | 0.158 |
| Continuous | 0.90 | (-0.09, 4.14) | 0.079 | 10.5 | (-7.38, 24.3) | 0.317 |
| **MONICA (replication)** |  |  |  |  |  |  |
| Cardiovascular endpoint |  |  |  |  |  |  |
| PWV $\geq 9$ vs $< 9$ m/s | 0.47 | (0.02, 1.41) | 0.028 | 4.85 | (-6.84, 12.13) | 0.238 |
| Continuous | 1.25 | (0.32, 2.44) | 0.004 | 3.42 | (-4.29, 13.03) | 0.232 |
| Total mortality |  |  |  |  |  |  |
| PWV $\geq 9$ vs $< 9$ m/s | 0.85 | (0.19, 1.94) | 0.002 | 6.94 | (-2.51, 14.31) | 0.108 |
| Continuous | 1.09 | (0.30, 2.04) | 0.006 | 0.02 | (-5.98, 8.73) | 0.707 |

Calculations were performed for the 5-year risk in IDCARS and the 10-year risk in MONICA. The base model included cohort (IDCARS only), sex, age (IDCARS) or age group (MONICA), mean arterial pressure, heart rate, body mass index, the total-to-HDL serum cholesterol ratio, smoking and drinking, diabetes and history of CV disease (IDCARS only). Pulse wave velocity (PWV) in MONICA was standardized to the subtraction method used in IDCARS. The integrated discrimination improvement (IDI) is the difference between the discrimination slopes of the base model and the base model extended by PWV. The discrimination slope is the difference in predicted probabilities (%) between participants without and with an endpoint. The net reclassification index (NRI) is the sum of the percentages of participants reclassified correctly in individuals without and with an endpoint (see Data Supplement pp **S8-S9**). IDI and NRI estimates are given with 95% CI.

### Rescaling of the PWV Threshold

In the analysis of the IDCARS and Copenhagen MONICA data the pulse wave travel distance was standardized to the subtraction method, as applied in IDCARS. To keep consistency with current guidelines and the software presently implemented in devices for PWV measurement, the 9-m/s threshold derived in IDCARS and replicated in MONICA was rescaled to account for the difference between the measured and anatomical pulse wave travel path. With this adjustment applied the 9-m/s threshold corresponded with 10 m/s (**Figure 2**).

**Figure 2.**
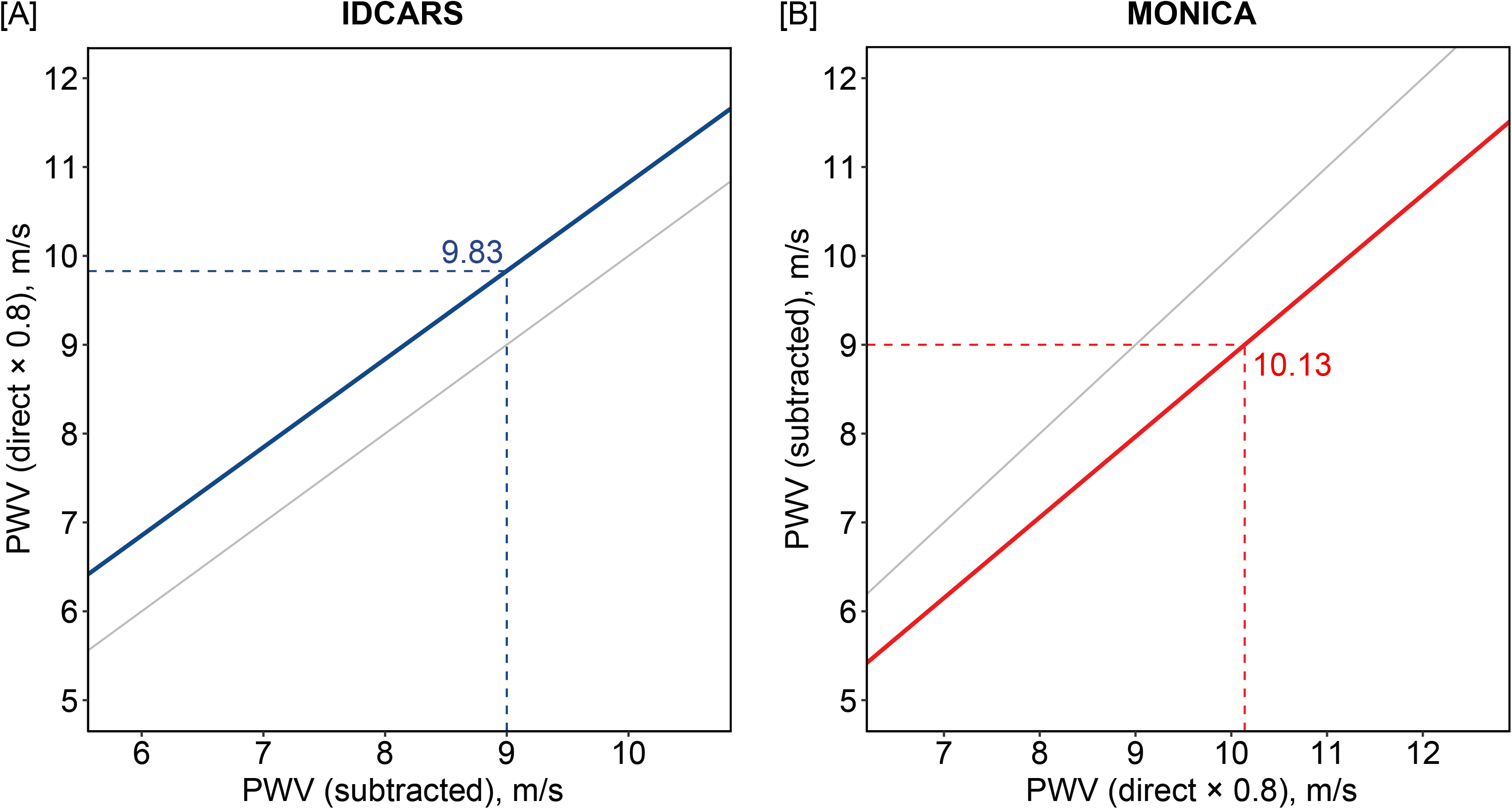
Rescaling the Outcome-Driven Pulse Wave Threshold for the Anatomical Pulse Wave Travel Distance Path In the analysis of the IDCARS and the Copenhagen MONICA data the pulse wave travel distance was standardized to the subtraction method, as applied in IDCARS. To keep consistency with current guidelines and clinical practice, the 9-m/s threshold derived in IDCARS and replicated in MONICA was rescaled to account for the difference between the measured and anatomical pulse wave travel path, using the formula published in references **2** and **7**. With this adjustment applied the 9-m/s threshold corresponded with 10 m/s. The gray line represents the line of identity.

## DISCUSSION

In IDCARS a two-pronged approach was applied to determine outcome-driven PWV thresholds in relation to the CV endpoint and total mortality. The PWV thresholds in well calibrated models converged to 9 m/s, of which the prognostic value was replicated in MONICA. The 2007 European Guideline for the Management of Hypertension^3^ proposed a risk-carrying PWV threshold of 12 m/s, because this level was believed to represent a rough estimate of high CV risk. The 2012 Consensus Document on the Measurement of PWV^2^ referred to the longitudinal patient and population studies published at that time to justify the 12-m/s threshold. However, the document^2^ went on stating that the 12-m/s cut-off limit was based on the direct measurement of the pulse transit distance. It therefore proposed a new standard distance, i.e., (common carotid artery – common femoral artery) × 0.8).^6^ Applying the new standard^6^ would result in a PWV threshold of 9.6 m/s, which was rounded to 10 m/s as an easy to remember value for use in daily clinical practice.^2^ In the current study, where relevant, the directly measured travel distance (MONICA) was converted to the subtraction distance (IDCARS)^2,7^ to increase comparability of the PWV estimates, either as descriptive variable (**Figure S1**) or as exposure variable. Accounting for the anatomical pulse wave travel distance showed that the 9-m/s threshold was equivalent with the 10-m/s cut-off, as proposed in the European guidelines.^3,7^

Several results presented in the current manuscript were generated as validation of the data resources being used. First, the sex distribution of anthropometric and hemodynamic characteristics and serum lipids was in line with the literature. In both IDCARS and MONICA, women compared with men had smaller body height, lower body weight and body mass index, lower systolic and diastolic BP and MAP, and higher HDL cholesterol (**Table S2**). Second, the main correlates of PWV were age, systolic BP, pulse pressure and MAP (**Table S4**). Third, to assess concordance with the literature, normal individuals were sampled from the IDCARS and the MONICA cohorts, using the same exclusion criteria as described by The Reference Values for Arterial Stiffness’ Collaboration (**Figure S2**).^7^ With standardization of the pulse wave travel path applied,^2,7^ PWV by age group was largely similar in IDCARS and MONICA and in agreement with the normal subgroup in the previously published cross-sectional meta-analysis (**Table S3**).

### Clinical Significance

Modelling time-to-event using proportional hazard regression implies that the association between adverse health outcomes and a risk factor is log-linear without a threshold at which the risk suddenly increases. Population studies of office^31^ or out-of-office^32^ BP or serum cholesterol^33^ have unmistakably illustrated this concept. Given that in the present analysis, the proportional hazard assumption for PWV was met, this construct is also applicable to PWV. For this reason, throughout the current manuscript, the risk with PWV was not only tabulated for the 9-m/s threshold, but also for a 1-SD increment in the continuously distributed PWV. Notwithstanding the continuous associations between adverse health outcomes and risk factors, operational or actionable thresholds of risk factors support clinicians in risk stratification and in identifying the need to start pharmacological treatment. Both in IDCARS and MONICA (**Table 3**), specificity of the 9-m/s threshold for the co-primary endpoints was ∼0.80, sensitivity was ∼0.55, and the overall predictive accuracy close to 0.75. IDI was significant for the CV endpoint in IDCARS and both co-primary endpoints in MONICA, whereas NRI did not reach statistical significance. The multivariable-adjusted IDI and NRI provide complementary information. Indeed, if addition of a marker to a model including several risk factors increases the predicted probability of an endpoint, this is reflected by a significant increase in IDI (**Table 4**). NRI indicates the extent to which a biomarker improves diagnostic accuracy, which in the current analyses was not significant, indicating that the discriminatory performance of PWV on top of commonly measured risk factors, in particular sex, age, various BP indexes (**Table S4**) and dyslipidemia, is small. A risk calculator is made available as **Data Supplement 2**. The SPARTE Investigators^34^ and the pathophysiology of aortic stiffness^35-37^ provide the interpretation of these findings. Aortic stiffness, as captured by PWV, integrates the lifetime injury to the arterial wall. Elastin and collagen are the major constituents of the extracellular matrix in the media of the central elastic arteries. Elastin provides reversible extensibility during systole, while collagen generates tensile strength. As people age, the elastin fibers become fragmented and the mechanical load is transferred to collagen fibers, which are up to 1000 times stiffer than elastin.^35^ This process already starts in young adulthood, but the deposition of elastin by vascular smooth muscle cells only occurs during fetal development and in early infancy, and is switched-off thereafter.^36^ This implies that elastin fiber damage is basically irreversible.^37^

In the SPARTE Trial,^34^ hypertensive patients were randomized to a therapeutic strategy targeting the normalization of PWV, measured every 6 months (N=264) or to a therapeutic strategy only implementing the European Hypertension Guidelines^3^ (N=272). After a median follow-up of 48.3 months, there was no significant between-group difference in the primary outcome, a composite CV endpoint (HR: 0.74; CI: 0.40-1.38). However, the secondary endpoints were met by showing that PWV-driven treatment for hypertension reduces office and ambulatory BP and aortic stiffening more than with application of BP-based guidelines. In a subgroup of 337 participants enrolled in the Systolic Blood Pressure Intervention Trial (SPRINT; 45% women; mean age: 64 years),^38^ intensive treatment (target systolic BP <120 mm Hg) compared with usual treatment (<140 mm Hg) produced a mean between-group reduction in systolic BP of 12.7 mm Hg (CI: 11.1-14.3 mm Hg) and at the end of the 18-month follow-up had attenuated the increase in PWV (9.0 *vs* 10 m/s; *P*<0.001). Basically, both trials highlighted the pathophysiological concept, that age and high BP are the main drivers of aortic stiffening. However, clinicians should be particularly concerned about patients, in whom there is disparity between PWV, age and MAP, and retrace the previous and current medical history of such patients to identify hidden risk factors. In the context of the current study, a PWV risk threshold of 9 m/s (or 10 m/s with the correction for the anatomical pulse wave travel path applied) should motivate clinicians to achieve stringent control of BP, in particular systolic BP, the extending force to be buffered by the elastin fibers.

### Limitations

The current study should be carefully interpreted within the context of its limitations. First, the diagnostic criteria and invasive management of coronary heart disease improved drastically from the early 1990s to the current state of the art. In MONICA only a single case of coronary revascularization was registered, whereas this number in IDCARS was 57 (**Table S5**). These period effects might explain why the 9-m/s PWV threshold was not replicated for coronary endpoints in MONICA, whereas PWV analyzed as continuously distributed variable retained significance (**Table S8**). Second, as shown by the NRI, the incremental value associated with PWV on top of all other risk factors was not significant (**Table 4**), explaining why the NRI results were not graphically translated into an analysis of the area under the curve of nested models. Finally, although IDCARS was a multi-ethnic cohort, Blacks were not represented in the current data resource.

### Perspectives

This individual-participants meta-analysis of longitudinal population studies with a composite CV endpoint and total mortality as co-primary endpoints identified, validated and replicated 9 m/s as new outcome-driven threshold for aortic PWV. With correction for the anatomical travel path, this cut-off corresponds with the 10-m/s threshold proposed in European guidelines.^3,7^ In quantitative terms, these outcome-driven thresholds refine risk stratification (IDI), albeit with a nonsignificant amount (NRI). In settings where PWV measurement can be implemented, exceeding the actionable thresholds should motivate clinicians to stringent management of modifiable CV risk factors, in particular systolic BP, which over a person’s lifetime leads to elastin fragmentation in the wall of the central arteries, thereby causing major CV complication and premature mortality.

## ARTICLE INFORMATION

### Sources of funding

The Non-Profit Research Association Alliance for the Promotion of Preventive Medicine, Mechelen, Belgium (www.appremed.org) received a nonbinding grant from OMRON Healthcare Co. Ltd., Kyoto, Japan, which supports the scholarships of D.-W.A, B.C., and Y.-L.Y. The grants which supported the cohort studies are listed by country.

*Argentina*: The Internal Medicine Service, Hospital Italiano de Buenos Aires, Buenos Aires, Argentina;

*Belgium*: European Union (HEALTH-F7-305507 HOMAGE), European Research Council (Advanced Researcher Grant 2011-294713-EPLORE and Proof-of-Concept Grant 713601-uPROPHET), European Research Area Net for Cardiovascular Diseases (JTC2017-046-PROACT), and Research Foundation Flanders, Ministry of the Flemish Community, Brussels, Belgium (G.0881.13);

*China*: The National Natural Science Foundation of China (grants 82270469, 82070432 and 82070435), the Ministry of Science and Technology (2018YFC1704902), Beijing, China, and by Shanghai Municipal Health Commission (2022LJ022 and 2017BR025);

*Czech Republic*: European Union (grants LSHM-CT-2006–037093 and HEALTH-F4-2007–201550) and Charles University Research program „Cooperatio – Cardiovascular Science“.

*Denmark*: 01-2-9-9A-22914 from the Danish Heart Foundation and R32-A2740 from the Lundbeck Fonden.

*Finland*: Academy of Finland (grant 321351), Emil Aaltonen Foundation, the Paavo Nurmi Foundation, the Urmas Pekkala Foundation, and the Hospital District of South-Western Finland;

*Italy*: European Union (grants LSHM-CT-2006–037093 and HEALTH-F4-2007–201550);

*Poland (Gdańsk)*: European Union (grants LSHM-CT-2006–037093 and HEALTH-F4-2007–201550);

*Poland (Kraków)*: European Union (grants LSHM-CT-2006–037093 and HEALTH-F4-2007–201550) and Foundation for Polish Science.

## Nonstandard Abbreviations and Acronyms

APPREMED: Non-Profit Research Association “*Alliance for the Promotion of Preventive Medicine*”
BP: blood pressure
CI: 95% confidence interval
CV: Cardiovascular
EPOGH: European Project on Genes in Hypertension
FLEMENGHO: Flemish Study on Environment Genes and Health Outcomes
HDL: high-density lipoprotein
HR: hazard ratio
IDCARS: International Database of Central Arterial Properties for Risk Stratification
IDI: integrated discrimination improvement
IQR: interquartile range
MONICA: Monitoring of Trends and Determinants in Cardiovascular Disease Health Survey – Copenhagen
MAP: Mean arterial pressure
NRI: net reclassification improvement
SPRINT: Systolic Blood Pressure Intervention Trial
PWV: aortic pulse wave velocity (carotid-femoral pulse wave velocity)

## Supplemental Material

Expanded Methods

Tables S1-S8

Figure S1-S5

## Novelty and Relevance

### What is New?

- IDCARS (N=3378) and MONICA (N=2458) are large population studies assuring generalizability.
- In well-calibrated multivariable models, the risk-carrying PWV thresholds in IDCARS converged to 9 m/s, of which the prognostic utility was replicated in MONICA.
- The 9-m/s PWV refined risk stratification on top of classical risk factors, albeit to a minor extent.
- Corrected for the anatomical pulse wave travel distance the 9-m/s threshold is equivalent to 10 m/s.

### What Are the Clinical Implications?

- Over a person’s lifetime, hypertension leads to irreparable elastin fragmentation in the wall of elastic arteries, thereby causing major CV complications and death.
- PWV integrates all unmodifiable and modifiable risk factors in a single variable, so that its measurement should be encouraged for risk stratification.
- Exceeding the risk-carrying PWV threshold should motivate clinicians to stringent management of risk factors, in particular hypertension.

